# Chest X-ray Severity and its Association with Outcomes in Patients with COVID-19 Presenting to the Emergency Department

**DOI:** 10.1101/2021.10.31.21265672

**Authors:** Daniel Kotok, Jose Rivera Robles, Christine Girard, Shruti Shettigar, Allen Lavina, Samantha Gillenwater, Andrew Kim, Anas Hadeh

**Affiliations:** Department of Pulmonary and Critical Care Medicine, Cleveland Clinic Florida, Weston, FL, USA; Department of Internal Medicine, Cleveland Clinic Florida, Weston, FL, USA

**Keywords:** Chest X-ray, Pulmonary Edema, COVID-19

## Abstract

**Background:** Severity of radiographic abnormalities on chest X-ray (CXR) in patients with COVID-19 has been shown to be associated with worse outcomes, but studies are limited by different scoring systems, sample size, patient age and study duration. Data regarding the longitudinal evolution of radiographic abnormalities and its association with outcomes is scarce. We sought to evaluate these questions using a well-validated scoring system (the Radiographic Assessment of Lung Edema [RALE] score) using data over 6 months from a large, multi-hospital healthcare system.

**Methods:** We collected clinical and demographic data and quantified radiographic edema on CXRs obtained in the emergency department (ED) as well as on days 1-2 and 3-5 (in those admitted) in patients with a nasopharyngeal swab positive for SARS-CoV-2 PCR visiting the ED for COVID-19-related complaints between March and September 2020. We examined the association of baseline and longitudinal evolution of radiographic edema with severity of hypoxemia and clinical outcomes.

**Results:** 870 patients were included (median age 53.6, 50.8% female). Inter-rate agreement for RALE scores was excellent (ICC = 0.84, 95% CI 0.82 - 0.87, p < 0.0001). RALE scores correlated with hypoxemia as quantified by SpO2-FiO2 ratio (r = -0.42, p < 0.001). Admitted patients had higher RALE scores than those discharged (6 [2, 11] vs 0 [0, 3], p < 0.001). An increase of RALE score of 4 or more was associated with worse 30-day survival (p < 0.01). Larger increases in the RALE score were associated with worse survival.

**Conclusions:** The RALE score is reproducible and easily implementable in adult patients presenting to the ED with COVID-19. Its association with physiologic parameters and outcomes at baseline and longitudinally makes it a readily available tool for prognostication and early ICU triage, particularly in patients with worsening radiographic edema.

## Introduction

The significant morbidity and mortality associated with the COVID-19 pandemic has resulted in expedited research and development of preventive, diagnostic and therapeutic strategies to decrease the incidence and severity of disease associated with SARS-CoV-2 (1, 2). The ability to provide accurate prognosis at time of diagnosis remains limited and relies mostly on baseline patient characteristics such as age and known co-morbidities (3–5). Combined radiographic, biomarker and artificial intelligence methods are emerging to assist with more accurate prognostication and identification of biologic phenotypes (6, 7).

Of the available diagnostic tools, plain chest radiography remains a simple and readily available tool that is accessible even in low-resource areas, some of which have been severely affected with extreme rates of infection and mortality(8). The Radiographic Assessment of Lung Edema has been shown to be accurate and reliable between observers with baseline assessment of CXR in patients with COVID-19 and non-COVID-19 acute respiratory distress syndrome (ARDS) (9–11). Although baseline radiographic edema has been shown to be associated with worse outcomes, whether the longitudinal evolution of radiographic edema as quantified by the RALE score is predicative of need for intensive care unit (ICU) admission, mechanical ventilation and 30-day survival remains largely unexplored.

We sought to independently evaluate the utility of the RALE score in a multi-center cohort of adult patients diagnosed with COVID-19 early in the pandemic and assess its association with severity of hypoxemia at baseline, to assess its ability to predict clinical outcomes at time of ED evaluation and in the early post-hospitalization period in those patients requiring hospital admission.

## Materials and Methods

### Patient Population

From March 2020 – October 2020, we collected data from the electronic medical records within the Cleveland Clinic system from 18 different clinical sites. We included all patients 18 years or older diagnosed with COVID-19 using nasal swab PCR. Patients with prior tracheostomy or those for whom the duration of symptoms associated with COVID-19 were unclear were excluded. Cleveland Clinic Institutional Review Board (IRB) exempted the study from IRB approval (FLA 20-038).

We collected baseline clinical data obtained at the time of emergency department (ED) visit (including demographics, comorbidities, physiologic and laboratory variables). We collected chest X-rays at time of ED evaluation. For patients admitted to the hospital, we also collected chest X-rays at two additional time periods: early (1-2 days) and late (3-5 days) after admission to the hospital. Fraction of inspired oxygen (FiO2) was calculated using 1 liter per minute (lpm) supplemental O2 to 3% FiO2 conversion and capped at 15 lpm O2. We examined the association of radiographic edema with hypoxemia, need for hospital admission, ICU admission, need for mechanical ventilation within 7 days of admission and 30-day mortality. For those admitted, we examined the association of worsening radiographic edema with 30-day survival and need for mechanical ventilation.

### RALE Score Calculation

We quantified the radiographic edema by calculating the RALE score as originally described by Warren and colleagues (9) Briefly, the RALE score assesses two aspects of pulmonary edema in four quadrants divided horizontally by a line through the first branch of the left main bronchus and vertically by the spinal column: *consolidation extent* (quantified as percent of CXR quadrant involved: 0% = 0; <25% = 1; 25-50% = 2; 50-75% = 3; >75% = 4) and *density* (1 = hazy; 2 = moderate; 3 = dense). Edema extent and density are multiplied for each quadrant and the total RALE score is obtained by addition of all quadrant scores (0-48) (Figure 1). Two independent reviewers (DK and SG) scored all CXRs.

**Figure 1.**
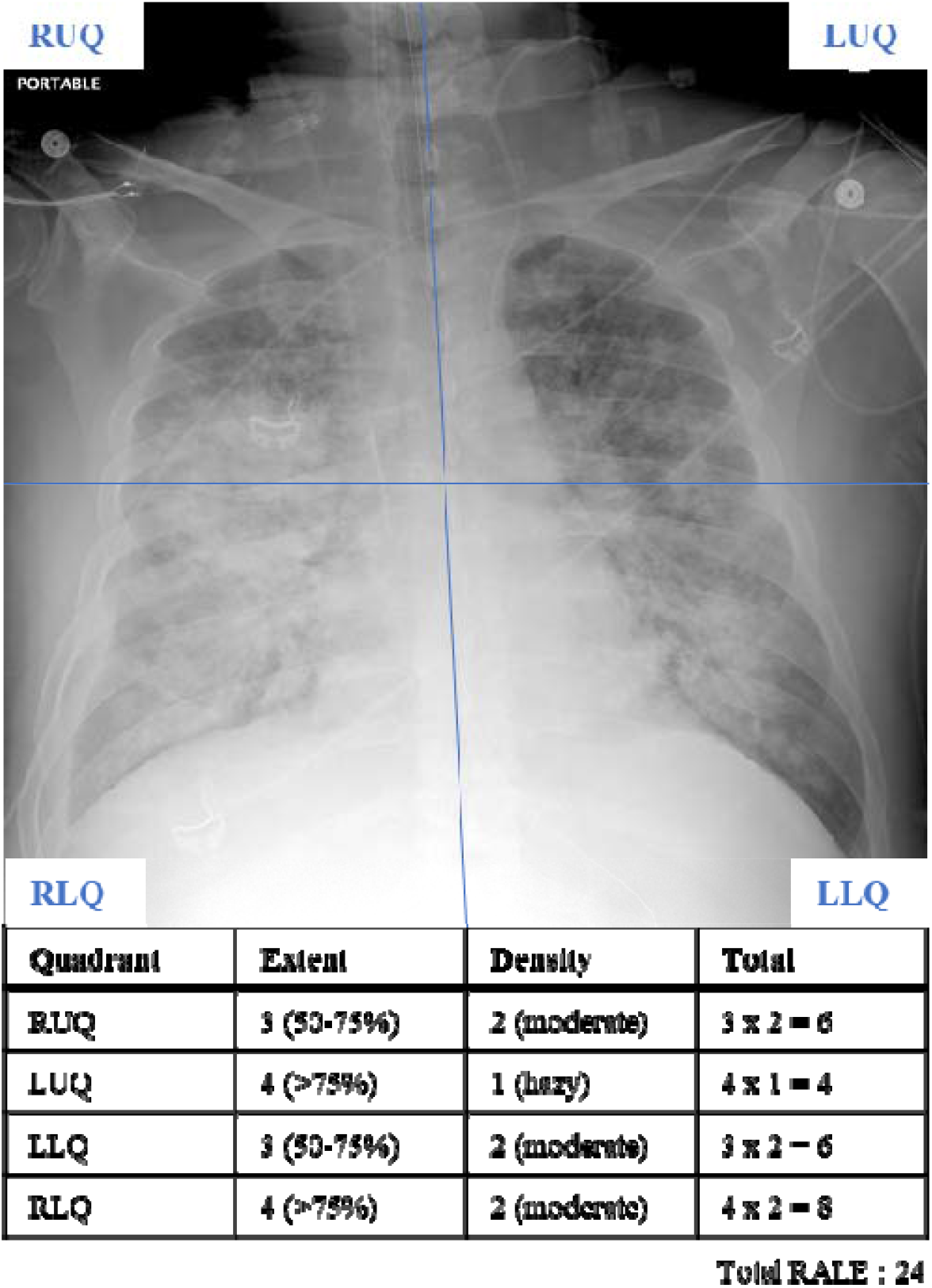
RALE score calculation from a chest radiograph (CXR). First, the CXR is divided in four quadrants defined by a drawing a horizontal line from the first branch of the left main bronchus and a vertical line through the spinal column separating the left and right lungs. Each quadrant is scored separately based on consolidation extent and density, which is then multiplied. The total RALE score is obtained by simple addition of all four quadrant scores (0-48).

### Statistical Analysis

We assessed interrater agreement (DK, SG) using the interclass correlation coefficient (ICC) with a two-way mixed agreement model (12, 13). We compared baseline characteristics using Wilcoxon signed-rank and Fisher’s exact tests for continuous and categorical variables, respectively. We applied Pearson’s correlation test to examine for baseline association between the RALE score and hypoxemia, i.e., S:F ratio (ratio of pulse oximetric saturation [SpO2] and fraction of inspired oxygen [F]). To derive the fraction of inspired oxygen, for every liter per minute (lpm) of inspired supplemental oxygen the fraction was increased by 0.03 (3%), capping at 15 lpm after which F was set to 1.

We divided the distribution of RALE score observations into four quartiles (Q1: 0 – 0, Q2: 1 – 2, Q3: 3 – 7, Q4: 8 – 32), and then examined associations of RALE quartiles with categorical clinical outcomes. We used Cox proportional hazard models adjusting for age, severity of hypoxemia and history of diabetes to examine the effects of baseline RALE scores on 30-day survival and need for intubation within 7 days of admission. To assess for the impact of radiographic edema worsening on outcomes, we considered different thresholds of RALE score increase from baseline CXR and then compared outcomes for patients with vs. without worsening edema. All statistical analyses were performed with the R statistical software, version 4.1 (14).

## Results

We reviewed the electronic medical records of 949 patients based on eligibility criteria. 21 patients did not have CXRs done on ED evaluation, while 58 patients had incomplete baseline and/or outcome data (Figure 2). A total of 870 patients were subsequently included in the study (Table 1). 376 (43%) patients had a normal CXR (RALE = 0). We further examined baseline characteristics of patients based on RALE quartiles (Table S1).

**Table 1.** Baseline characteristics of included subjects at time of ED evaluation. BMI: Body mass index; S:F Ratio: Ratio of pulse oximetric oxygen saturation and fraction of inspired oxygen; RALE Score: Radiographic assessment of lung edema score.

|  |  |
| --- | --- |
| n | 870 |
| Male (%) | 428 (49.2) |
| Age, years (median [IQR]) | 53.60 [39.0, 66.3] |
| BMI (median [IQR]) | 30.53 [26, 35.6] |
| Symptom duration (median [IQR]) | 4.0 [2.0, 7.0] |
| History of diabetes (%) | 183 (21.1) |
| History of tobacco use (%) | 318 (37.0) |
| SF Ratio (median [IQR]) | 461.90 [452.4, 471.4] |
| RALE score (median [IQR]) | 2.00 [0, 7.0] |
| 30-day Mortality (%) | 65 (7.5) |

**Figure 2.**
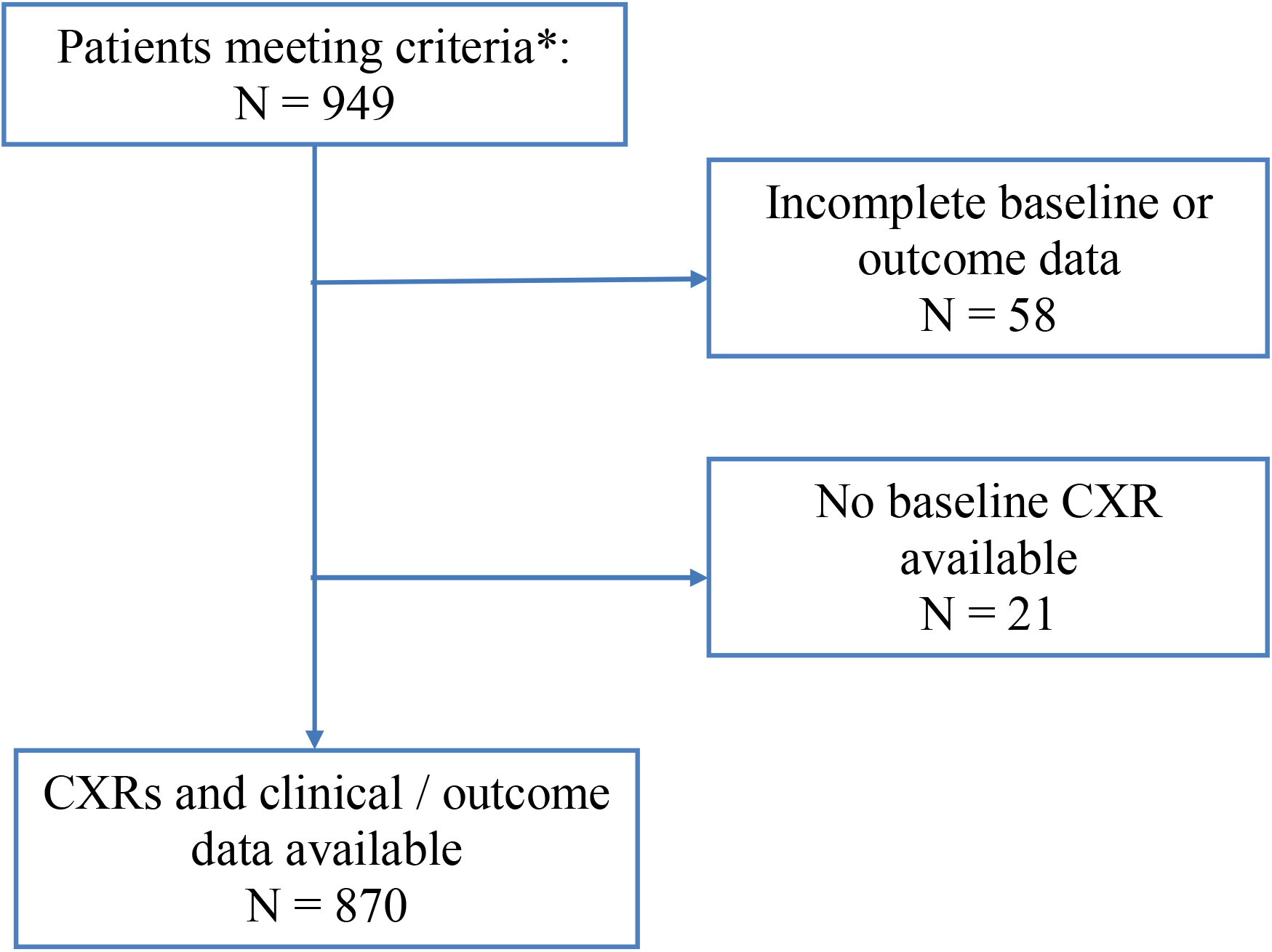
Selection process of patients. We identified 949 patients based on eligibility criteria. 58 patients did not have baseline demographic or outcome data available. 21 patients did not have baseline CXRs. A total of 870 patients had both CXRs and data required for analysis. * We included patients 18 years or older diagnosed with COVID-19 using nasal swab PCR. Patients with prior tracheostomy or those for whom the clinical course and duration of symptoms associated with COVID-19 were unclear were excluded.

### Inter-reviewer Agreement for RALE Score

A total of 1185 of CXRs were reviewed from different study periods. Inter-rate agreement between the two reviewing clinicians (DK, SG) for the RALE score was good (interclass correlation coefficient = 0.84, 95% CI 0.82 - 0.87, p < 0.001).

### Association with baseline demographic and clinical variables

Patients in the higher RALE quartiles were more likely to be male, older, have history of obstructive sleep apnea, heart failure, hypertension, hyperlipidemia, chronic kidney disease, lung disease, diabetes and immunosuppression (p < 0.05, Table S1) but not tobacco use (p = 0.28). Ethanol consumption was less common in the higher RALE quartiles (p = 0.002). They were also more likely to be tachypneic, have lower hemoglobin levels and higher neutrophil-to-lymphocyte ratios. At baseline, there was a significant association between severity of hypoxemia and RALE score (Pearson’s r = -0.42, p < 0.001, Figure S1).

### Association with clinical outcomes

The RALE score was significantly higher in patients requiring hospitalization (2 [6-11]) compared to those who were discharged home (0 [0-3], p < 0.001) (Figure 3A). In those patients requiring admission, a higher RALE scored was observed in those requiring ICU admission (4 [8-14)) compared to those that did not (5 [0-9], p < 0.001) (Figure 3B). A higher baseline RALE score was associated with unadjusted 30-day mortality (5 [8-15] vs 0 [1-6], p < 0.001) and need for mechanical ventilation (8 [5-16] vs 0 [1-6], p < 0.001). In a multivariate Cox-proportional hazards model adjusted for age, severity of hypoxemia and history of diabetes, we found baseline RALE scores to not be associated with 30-day survival when the first two quartiles were compared (HR 1.5 [0.13 – 16.6], p = 0.74). However, we found 30-day survival to be significantly lower in the 3^rd^ the 4^th^ quartiles when compared to the 1^st^ RALE quartile (HR 11.6 [2.7 – 49.3], p < 0.001 and 10.1 [2.3 – 43], p = 0.002, respectively) (Figure 4). Similar results were observed with regards to the need for intubation and mechanical ventilation within 7 days of admission (Q3 vs Q1 and Q4 vs Q1 – HR 6.49 [1.86 – 22.54] and HR 9.48 [2.76 – 32.56], p = 0.003 and p = 0.0003, respectively) (Figure S2).

**Figure 3.**
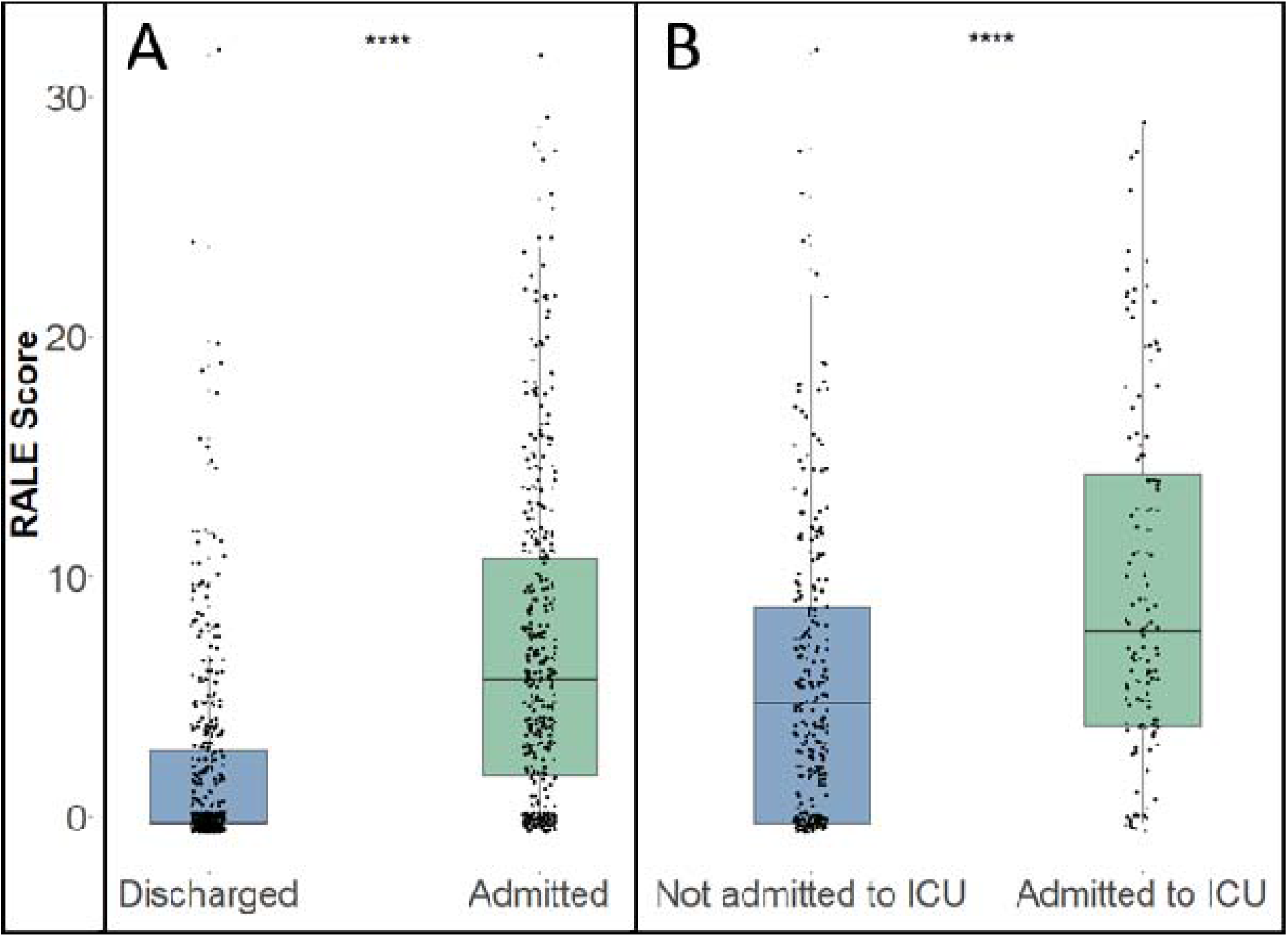
Comparison of median RALE score by need for hospital and ICU admission. Patients requiring hospital admission had a higher median RALE score compared to those discharged home (p < 0.001) (Panel A). Similarly, in patients admitted to the hospital – those who required ICU admission had a higher median RALE score compared to those who did not (Panel B) (p < 0.001).

**Figure 4.**
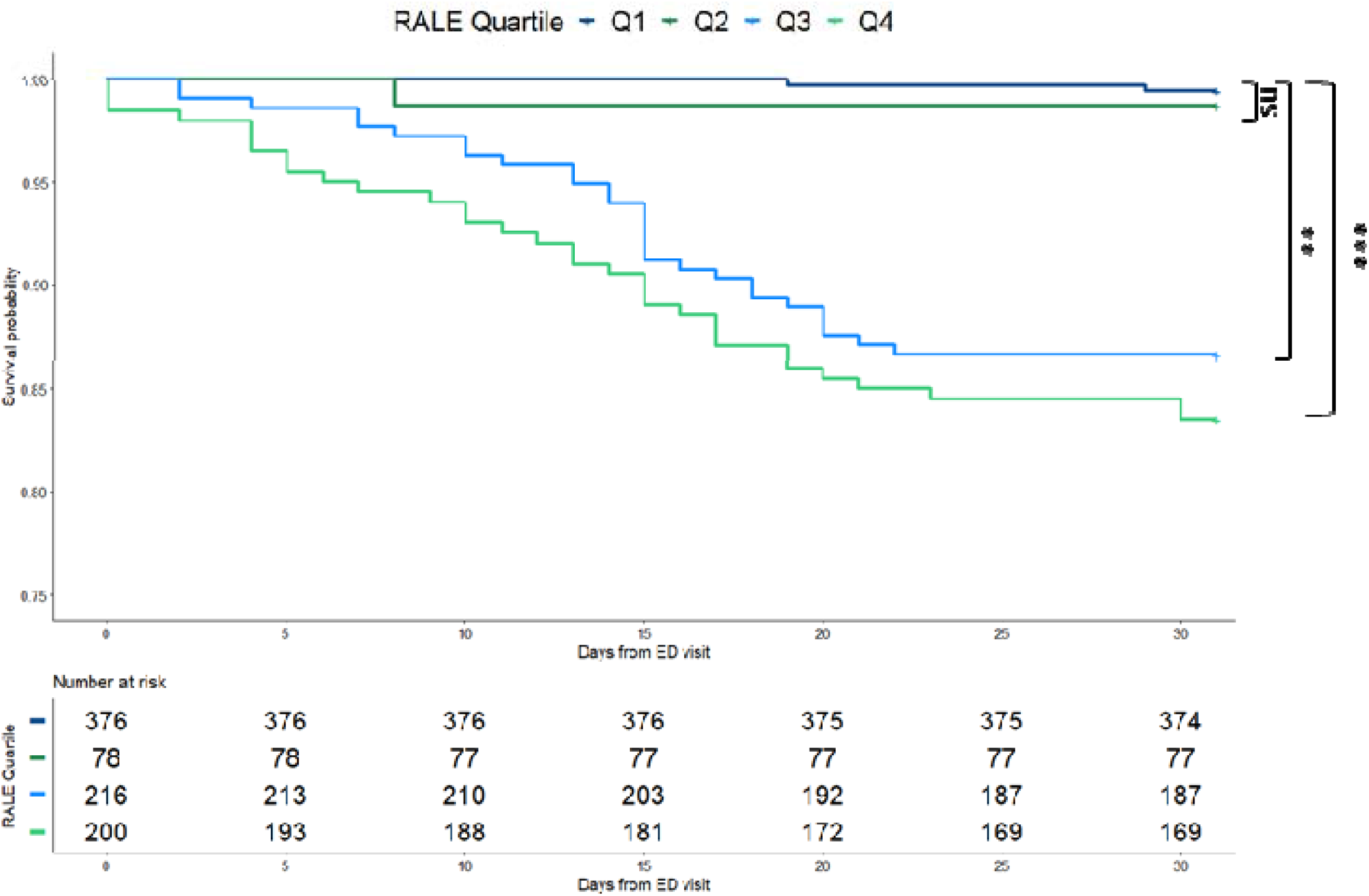
Kaplan-Meier estimates of 30-day survival comparing RALE score quartiles. We observed significantly lower likelihood of survival at 30 days when Q3 and Q4 were compared to Q1 (** p = 0.002 and **** p < 0.001, respectively). There was no significant difference in survival when Q1 and Q2 were compared.

**Figure 5.**
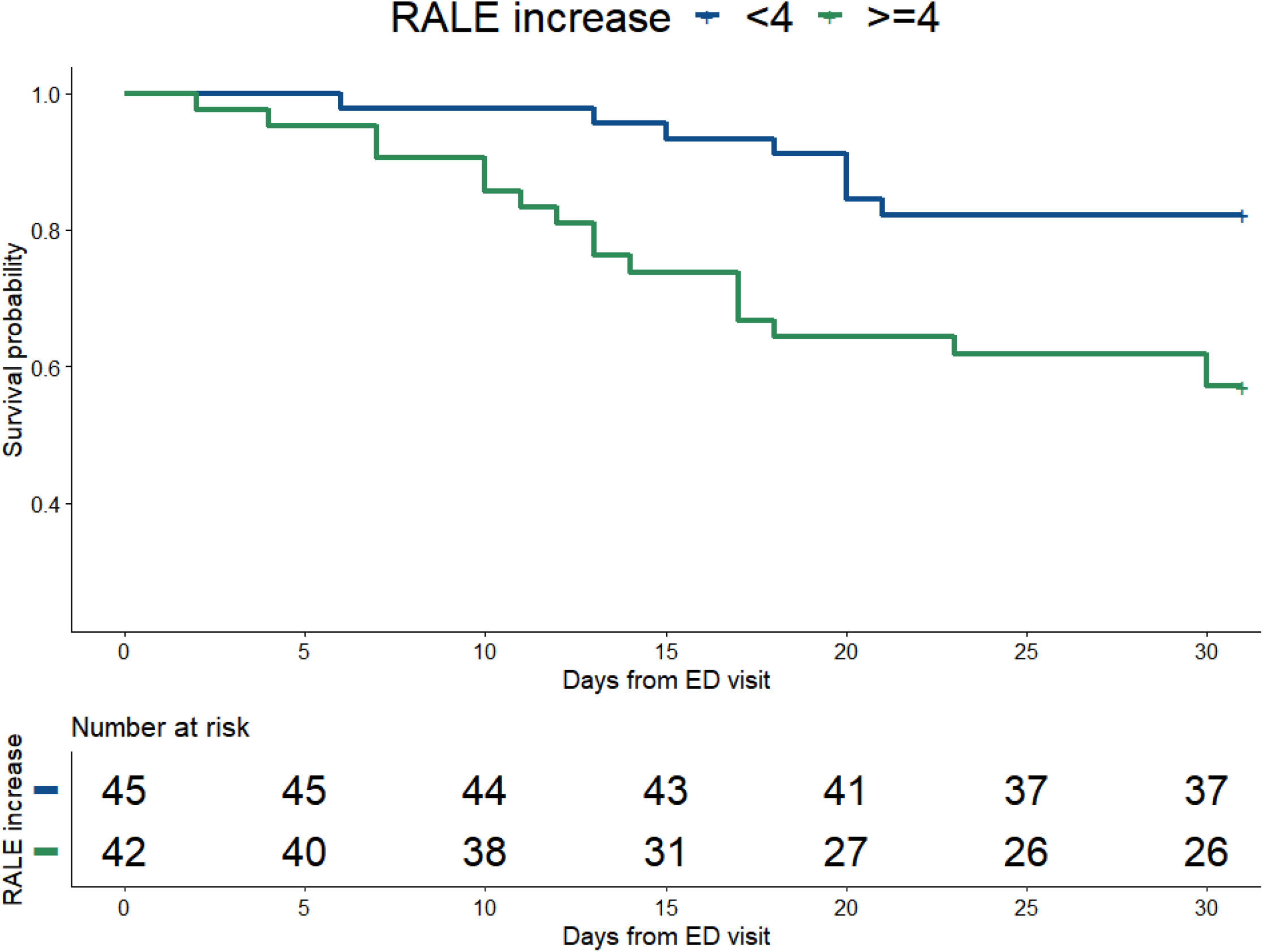
Kaplan-Meier estimates of 30-day survival based on the longitudinal evolution of RALE score over time. A 4-point increase in RALE score or more from baseline to early period was associated with worse 30-day survival (p = 0.002).

### Longitudinal evolution of radiographic edema in hospitalized patient and its association with outcomes

Among hospitalized patients, 408 had baseline, 91 had early-period and 123 had late-period CXRs. There were significant differences between baseline and early period (6 [2-11] vs 14 [5-20], p < 0.001) as well as baseline and late (6 [2-11] vs 11 [5-20], p < 0.001) period RALE scores. There was no difference between the early and late period (p = 0.51) (Figure S3). A 4-point increase in RALE score or more from baseline to early period was associated with worse 30-day survival (HR 2.1 [1-2 – 3.5], p = 0.006) (Figure 4). This was also true for increase from baseline to late (HR 2.7 [1.7 -4.2], p = 0.002. Further increase in the RALE score was associated with worse survival (Table 2). Although we did not observe identical trends in need for mechanical ventilation, a larger increase in the RALE score from baseline to early (>=6) or a later increase in the RALE score of >=4 (baseline to late) was associated with higher likelihood of requiring mechanical ventilation (HR 2.2 [1.1 – 4.3], p = 0.02 and HR 6.1 [3.4 – 10.7], p < 0.0001, respectively) (Table 2).

**Table 2.** Cox regression analysis for progressive increases in RALE score, 30-day survival, and 7-day probability of not requiring mechanical ventilation. A RALE score increases of at least 4 points from baseline to either early or late periods was associated with worse 30-day survival. A larger early increase (baseline to early by 6 or more) or a later (baseline to late by 4 or more) increase in the RALE score was associated with higher probability of requiring mechanical ventilation. All analyses were done after adjustment for baseline hypoxemia, age and history of diabetes.

| Examined parameter | RALE score increase<br>(>=) | Hazard ratio [95% CI] | p value |
| --- | --- | --- | --- |
| **Likelihood of mortality at 30 days** |  |  |  |
| Baseline to early | 4 | 2.07 [1.23 - 3.49] | **0.006** |
|  | 5 | 2.26 [1.35 - 3.79] | **0.002** |
|  | 6 | 3.06 [1.81 - 5.16] | **< 0.001** |
|  | 7 | 3.58 [2.08 - 6.14] | **< 0.001** |
|  | 8 | 3.87 [2.26 - 6.63] | **< 0.001** |
| Baseline to late | 4 | 2.67 [1.69 - 4.22] | **< 0.001** |
|  | 5 | 2.57 [1.63 - 4.04] | **< 0.001** |
|  | 6 | 2.65 [1.69 - 4.16] | **< 0.001** |
|  | 7 | 2.83 [1.8 - 4.46] | **< 0.001** |
|  | 8 | 2.9 [1.85 - 4.55] | **< 0.001** |
| Early to late | 4 | 0.97 [0.36 - 2.65] | 0.958 |
|  | 5 | 0.58 [0.18 - 1.89] | 0.364 |
|  | 6 | 0.62 [0.18 - 2.13] | 0.451 |
|  | 7 | 1.20 [0.38 - 3.82] | 0.76 |
|  | 8 | 0.98 [0.29 - 3.37] | 0.979 |
| **Likelihood of requiring intubation at 7 days** |  |  |  |
| Baseline to early | 4 | 1.58 [0.82 - 3.04] | 0.171 |
|  | 5 | 1.79 [0.93 - 3.46] | 0.082 |
|  | 6 | 2.20 [1.14 - 4.27] | **0.02** |
|  | 7 | 2.38 [1.21 - 4.69] | **0.012** |
|  | 8 | 1.99 [1.04 - 3.8] | **0.037** |
| Baseline to late | 4 | 6.06 [3.44 - 10.67] | **< 0.001** |
|  | 5 | 5.59 [3.26 - 9.56] | **< 0.001** |
|  | 6 | 5.89 [3.45 - 10.07] | **< 0.001** |
|  | 7 | 6.61 [3.89 - 11.26] | **< 0.001** |
|  | 8 | 6.85 [4.06 - 11.57] | **< 0.001** |
| Early to late | 4 | 0.79 [0.37 - 1.67] | 0.534 |
|  | 5 | 0.64 [0.28 - 1.46] | 0.289 |
|  | 6 | 0.76 [0.33 - 1.78] | 0.532 |
|  | 7 | 0.69 [0.28 - 1.67] | 0.406 |
|  | 8 | 0.59 [0.22 - 1.55] | 0.283 |

## Discussion

In this multicenter, retrospective, broadly inclusive study of 870 patients presenting to the emergency department with symptoms concerning for and diagnosed with COVID-19 using nasal-swab PCR between March and September 2020, we examined the association of radiographic edema on CXRs using the RALE score with physiologic parameters and clinical outcomes. Patients with worse radiographic edema as quantified by the RALE score on admission were more likely to be hypoxemic, require hospital admission, ICU admission, mechanical ventilation, and higher 30-day mortality. In addition to these baseline associations, we were able to demonstrate that progressive worsening of radiographic edema over the course of hospitalization for those requiring admission conferred a higher risk of death and need for intubation. This risk increased with worsening of radiographic edema even after adjustment for variables commonly associated with worse outcomes in patients with COVID-19 such as history of diabetes, age, and baseline hypoxemia.

The association between baseline radiographic edema and worse outcomes has been consistently demonstrated globally in a multitude of smaller studies using several scoring scores demonstrating an association between CXR severity and clinical outcomes (15–21). Our study is the largest to date utilizing the RALE score, which has been previously shown to have high rates of inter-observer agreement across different diseases including pneumonia, ARDS and heart failure (10, 11, 21). In addition to re-demonstrating previously reported data (i.e. worse radiographic edema being associated with worse outcomes) we were also able to demonstrate a very low-mortality in those with minimal radiographic edema (Q1 and Q2 with RALE score of 2 or less). This findings further reinforces the notion that screening CXRs in patients with asymptomatic or mild COVID-19 disease is not required (22). However, if performed, a normal CXR at time of ED visit is an indicator of a favorable prognosis. The ability of CXR to identify patients who are at low risk for progression to severe disease while also providing prognostic value for those with moderate-severe (Q3 and Q4 RALE score of 3 or more) radiographic edema provides a readily available tool at time of ED evaluation. Combined with deep-learning, radiographic evaluation using CXRs in COVID-19 has recently been shown to have an even higher predictive value compared to CXR or clinical data alone (7).

Although baseline radiographic edema has been repeatedly shown to be of predictive value, the value of longitudinal evaluation using CXR shortly after admission remains largely unclear. Small studies examining the evolution of CT abnormalities showed mixed results. This was likely due to a combination of heterogeneity in disease severity and timing of CT imaging (16, 23). Studies examining the association of baseline and longitudinal evolution of radiographic edema on CXR in patients with ARDS showed mixed results. These outcomes are presumably due to non-pulmonary ARDS affecting outcomes and the variability in agreement between clinicians with regards to what is labeled as ARDS (11, 24, 25). In the case of COVID-19 (with or without ARDS), where initial pathology requiring hospital admission is often confined to the lungs, we hypothesized that tracking radiographic edema may provide prognostic information outside of vital and clinical monitoring. The significant association between worsening radiographic edema with worse outcomes even after adjustments for confounding factors suggests that progression of COVID-19 pulmonary disease is adequately captured on CXR and periodic radiography may assist in prognostication in hospitalized patients with COVID-19.

There are several limitations associated with our study. The time-frame during which the study was performed did not allow for adjustment for anti-viral treatments (such as dexamethasone and remdesivir) which have been shown to decrease severity of radiographic edema (26) as these were introduced later during the study period within the healthcare system (late August-September). We were also not able to adjust for any variables other than those obtained at baseline when performing the longitudinal analyses, raising the question of whether other organ failure outside of the pulmonary system (renal) and fluid status have contributed to radiographic deterioration that is not directly associated with COVID-19 pulmonary disease. Lastly, while the study is a multi-center study across multiple hospitals and states in the United States, practices and procedures early in the pandemic were often institution-based and not necessarily guided by the Centers for Disease Control and Prevention and/or the World Health Organization. As such, institutional standards of treatment may not have been reflective of other healthcare systems around the same time period. Despite these limitations, we excluded patients for which there was incomplete data that would otherwise require imputation and potentially affect our analyses. Instead, the study protocol structured the CXR sampling periods to provide early, standardized meaningful clinical data that may assist clinicians in prognostication and decision making in patients with COVID-19.

## Conclusion

In conclusion, we were able to demonstrate the pragmastim and reproducibility of the RALE score in patients with COVID-19 which showed significant associations between severity of radiographic edema and clinical outcomes both at baseline and longitudinally in patients hospitalized with COVID-19. The degree of progression as dictated by the RALE score was linked to worse outcomes. Furthermore, our study demonstrates that longitudinal assessment of radiographic edema may provide prognostic value and assist clinicians in prognostication in patients with COVID-19. The effect of antiviral therapy on these findings is unclear and requires further investigation.

## Data Availability

All data produced in the present study are available upon reasonable request to the authors

## List of abbreviations

ARDS: Acute respiratory distress syndrome
RALE: Radiographic Assessment of Lung Edema
CXR: Chest X-ray
COVID-19: Coronavirus disease 2019
ED: Emergency department
ICU: Intensive care unit
ICC: Interclass correlation coefficient
S:F ratio: Ratio of pulse oximetric saturation [SpO2] and fraction of inspired oxygen [F]
Q: Quartile

## Contributions

**Daniel Kotok:** Guarantor of the Paper, Investigation, Software, Formal analysis, Writing - Original Draft, Writing - Review & Editing, Visualization

**Jose Rivera Robles:** Investigation, Writing - Review & Editing

**Christine Girard:** Investigation, Writing - Review & Editing

**Shruti Shettigar:** Investigation, Writing - Review & Editing

**Allen Lavina:** Investigation, Writing - Review & Editing

**Samantha Gillenwater:** Investigation, Writing - Review & Editing

**Andrew Kim:** Conceptualization, Methodology, Investigation, Writing - Original Draft, Writing - Review & Editing, Supervision

**Anas Hadeh:** Conceptualization, Methodology, Investigation, Writing - Original Draft, Writing - Review & Editing, Supervision

## References

1. Xie J, Tong Z, Guan X, Du B, Qiu H, Slutsky AS. Critical care crisis and some recommendations during the COVID-19 epidemic in China. Intensive Care Med 2020;46:837–840.

2. Douillet D, Penaloza A, Mahieu R, Morin F, Chauvin A, Gennai S, Schotte T, Montassier E, Thiebaud P-C, François AG, Dall’acqua D, Benhammouda K, Bissokele P, Violeau M, Joly L-M, Andrianjafy H, Soulie C, Savary D, Riou J, Roy P-M, Andrianjafy H, Baudin L, Benhammouda K, Bissolokele P, Brice C, Cayeux C, Casalino E, Casarin C, Chauvin A, et al. Outpatient Management of Patients With COVID-19: Multicenter Prospective Validation of the Hospitalization or Outpatient Management of Patients With SARS-CoV-2 Infection Rule to Discharge Patients Safely. Chest 2021;160:1222–1231.

3. Du N, Jiang Y, Zhang Q, Che L, Li X, Lou L, Bao W, Hua S. Clinical characteristics of family-clustered onset of coronavirus disease 2019 in Jilin Province, China. https://doi.org/101080/2150559420201816075 2020;11:1240–1249.

4. Xie J, Covassin N, Fan Z, Singh P, Gao W, Li G, Kara T, Somers VK. Association Between Hypoxemia and Mortality in Patients With COVID-19. Mayo Clin Proc 2020;95:1138.

5. Mo P, Xing Y, Xiao Y, Deng L, Zhao Q, Wang H, Xiong Y, Cheng Z, Gao S, Liang K, Luo M, Chen T, Song S, Ma Z, Chen X, Zheng R, Cao Q, Wang F, Zhang Y. Clinical characteristics of refractory COVID-19 pneumonia in Wuhan, China. Clin Infect Dis 2020; doi:10.1093/cid/ciaa270.

6. Drohan CM, Nouraie SM, Bain W, Shah FA, Evankovich J, Zhang Y, Morris A, McVerry BJ, Kitsios GD. Biomarker-Based Classification of Patients With Acute Respiratory Failure Into Inflammatory Subphenotypes: A Single-Center Exploratory Study. Crit Care Explor 2021;3:e0518.

7. Jiao Z, Choi JW, Halsey K, Tran TML, Hsieh B, Wang D, Eweje F, Wang R, Chang K, Wu J, Collins SA, Yi TY, Delworth AT, Liu T, Healey TT, Lu S, Wang J, Feng X, Atalay MK, Yang L, Feldman M, Zhang PJL, Liao W-H, Fan Y, Bai HX. Prognostication of patients with COVID-19 using artificial intelligence based on chest x-rays and clinical data: a retro1. Jiao Z et al. Prognostication of patients with COVID-19 using artificial intelligence based on chest x-rays and clinical data: a ret. Lancet Digit Heal 2021;3:e286–e294.

8. Lancet T. India’s COVID-19 emergency. Lancet (London, England) 2021;397:1683.

9. Warren MA, Zhao Z, Koyama T, Bastarache JA, Shaver CM, Semler MW, Rice TW, Matthay MA, Calfee CS, Ware LB. Severity scoring of lung oedema on the chest radiograph is associated with clinical outcomes in ARDS. Thorax 2018;73:840–846.

10. Kotok D, Yang L, Evankovich JW, Bain W, Dunlap DG, Shah F, Zhang Y, Manatakis DV, Benos PV, Barbash IJ, Rapport SF, Lee JS, Morris A, McVerry BJ, Kitsios GD. The evolution of radiographic edema in ARDS and its association with clinical outcomes: A prospective cohort study in adult patients. J Crit Care 2020;56:.

11. M J, J A, B P, S J, JY L, R B, T G, E F, C L, JE B, JA B, JM C, LB W. Early Changes Over Time in the Radiographic Assessment of Lung Edema Score Are Associated With Survival in ARDS. Chest 2020;158:2394–2403.

12. McGraw KO, Wong SP. Forming inferences about some intraclass correlation coefficients. Psychol Methods 1996;1:30–46.

13. Bartko JJ. The Intraclass Correlation Coefficient as a Measure of Reliability. Psychol Rep 1966;19:3–11.

14. R: The R Project for Statistical Computing. at <https://www.r-project.org/>.

15. Booth A, Reed AB, Ponzo S, Yassaee A, Aral M, Plans D, Labrique A, Mohan D. Population risk factors for severe disease and mortality in COVID-19: A global systematic review and meta-analysis. PLoS One 2021;16:e0247461.

16. Liu J, Chen T, Yang H, Cai Y, Yu Q, Chen J, Chen Z, Shang Q-L, Ma C, Chen X, Xiao E. Clinical and radiological changes of hospitalised patients with COVID-19 pneumonia from disease onset to acute exacerbation: a multicentre paired cohort study. Eur Radiol 2020 3010 2020;30:5702–5708.

17. Borghesi A, Zigliani A, Golemi S, Carapella N, Maculotti P, Farina D, Maroldi R. Chest X-ray severity index as a predictor of in-hospital mortality in coronavirus disease 2019: A study of 302 patients from Italy. Int J Infect Dis 2020;96:291–293.

18. Monaco CG, Zaottini F, Schiaffino S, Villa A, Pepa G Della, Carbonaro LA, Menicagli L, Cozzi A, Carriero S, Arpaia F, Leo G Di, Astengo D, Rosenberg I, Sardanelli F. Chest x-ray severity score in COVID-19 patients on emergency department admission: a two-centre study. Eur Radiol Exp 2020;4:.

19. Balbi M, Caroli A, Corsi A, Milanese G, Surace A, Di Marco F, Novelli L, Silva M, Lorini FL, Duca A, Cosentini R, Sverzellati N, Bonaffini PA, Sironi S. Chest X-ray for predicting mortality and the need for ventilatory support in COVID-19 patients presenting to the emergency department. Eur Radiol 2020;1–14. doi:10.1007/s00330-020-07270-1.

20. Gatti M, Calandri M, Barba M, Biondo A, Geninatti C, Gentile S, Greco M, Morrone V, Piatti C, Santonocito A, Varello S, Bergamasco L, Cavallo R, Di Stefano R, Riccardini F, Boccuzzi A, Limerutti G, Veltri A, Fonio P, Faletti R. Baseline chest X-ray in coronavirus disease 19 (COVID-19) patients: association with clinical and laboratory data. Radiol Medica 2020;125:1271–1279.

21. Cozzi D, Albanesi M, Cavigli E, Moroni C, Bindi A, Luvarà S, Lucarini S, Busoni S, Mazzoni LN, Miele V. Chest X-ray in new Coronavirus Disease 2019 (COVID-19) infection: findings and correlation with clinical outcome. Radiol Medica 2020;125:730–737.

22. Kuo BJ, Lai YK, Tan MLM, Goh X-YC. Utility of Screening Chest Radiographs in Patients with Asymptomatic or Minimally Symptomatic COVID-19 in Singapore. https://doi.org/101148/radiol2020203496 2020;298:E131–E140.

23. Gao Y, Hu Y, Zhu J, Liu H, Qiu R, Lin Q, He X, Lin H Bin, Cheng S, Li G. The value of repeated CT in monitoring the disease progression in moderate COVID-19 pneumonia: A single-center, retrospective study. Medicine (Baltimore) 2021;100:e25005.

24. Kotok D, Yang L, Evankovich JW, Bain W, Dunlap DG, Shah F, Zhang Y, Manatakis D V., Benos P V., Barbash IJ, Rapport SF, Lee JS, Morris A, McVerry BJ, Kitsios GD. The evolution of radiographic edema in ARDS and its association with clinical outcomes: A prospective cohort study in adult patients. J Crit Care 2020; doi:10.1016/j.jcrc.2020.01.012.

25. Figueroa-Casas JB, Brunner N, Dwivedi AK, Ayyappan AP. Accuracy of the chest radiograph to identify bilateral pulmonary infiltrates consistent with the diagnosis of acute respiratory distress syndrome using computed tomography as reference standard. J Crit Care 2013;28:352–357.

26. Kim W-Y, Kweon OJ, Cha MJ, Baek MS, Choi S-H. Dexamethasone may improve severe COVID-19 via ameliorating endothelial injury and inflammation: A preliminary pilot study. PLoS One 2021;16:e0254167.

