## Supplemental for "Chest X-ray Severity and its Association with Outcomes in Patients with COVID-19 Presenting to the Emergency Department"

### Supplement

**Supplemental figures: pages 2-4**

**Supplemental table: pages 5-6**

### Supplemental Figure 1. Association between SF ratio and RALE score. There was a significant, negative correlation between the RALE score and hypoxemia as quantified by the SF ratio (Pearson’s r = - 0.42, p < 0.001).

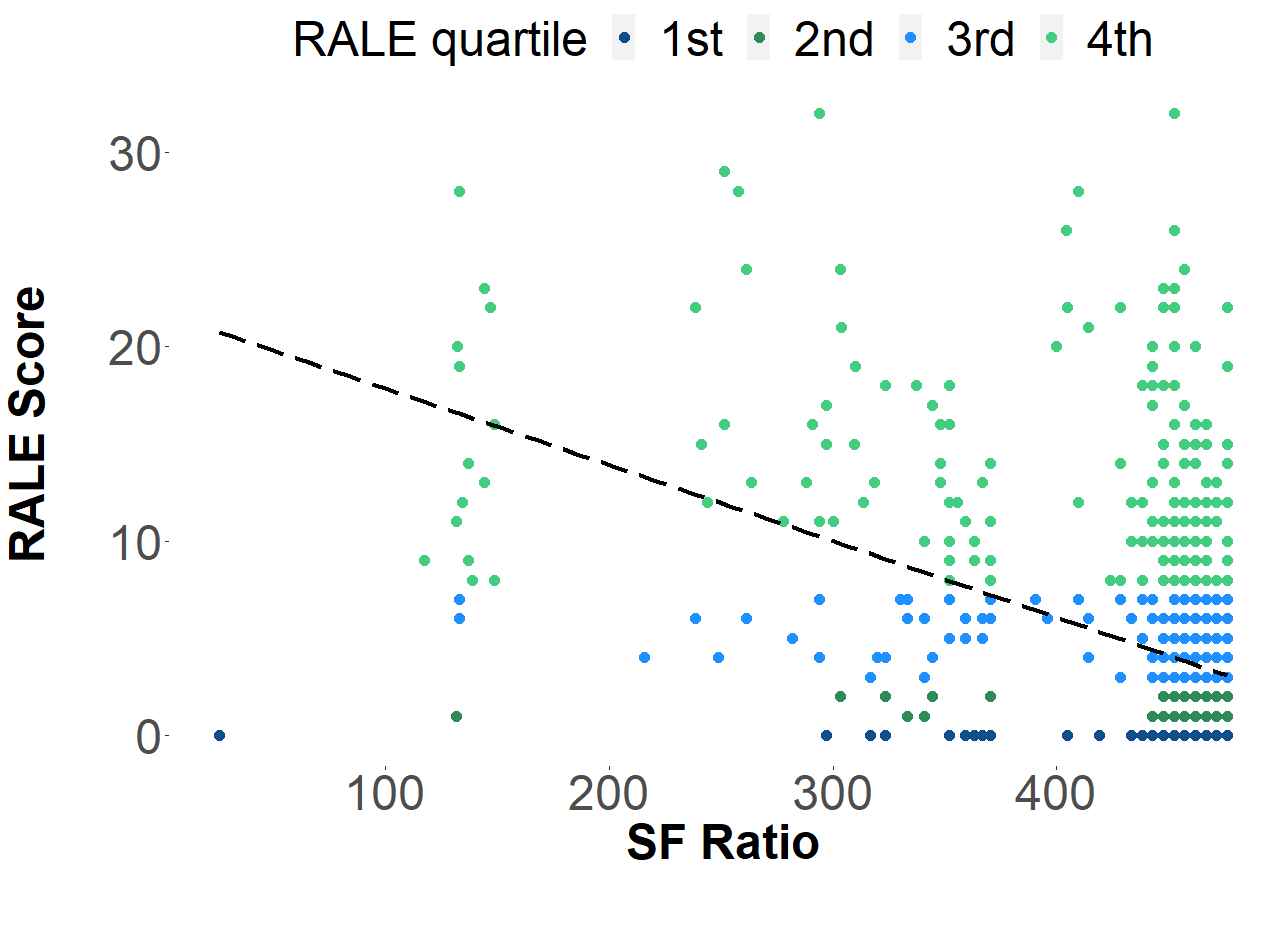

**Supplemental Figure 2. Kaplan-Meier estimates of 7-day probability of not requiring mechanical ventilation comparing RALE score quartiles.** We observed significantly lower likelihood of not requiring mechanical ventilation at 7 days when Q1 was compared with Q3 and Q4. There was no significant difference when Q1 and Q2 were compared.

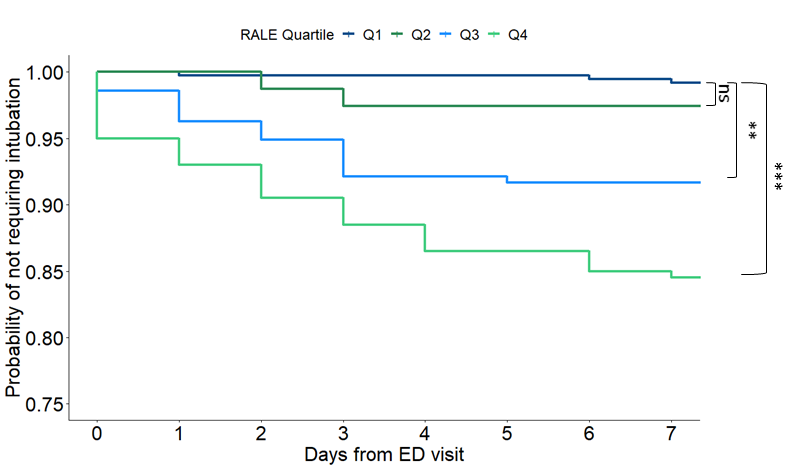

**Supplemental Figure 3. Median RALE comparing different study periods.** There was no significant difference between the median RALE score when the early and late periods were compared. When compared to baseline RALE scores, however, both periods had significantly higher median RALE scores (p < 0.001).

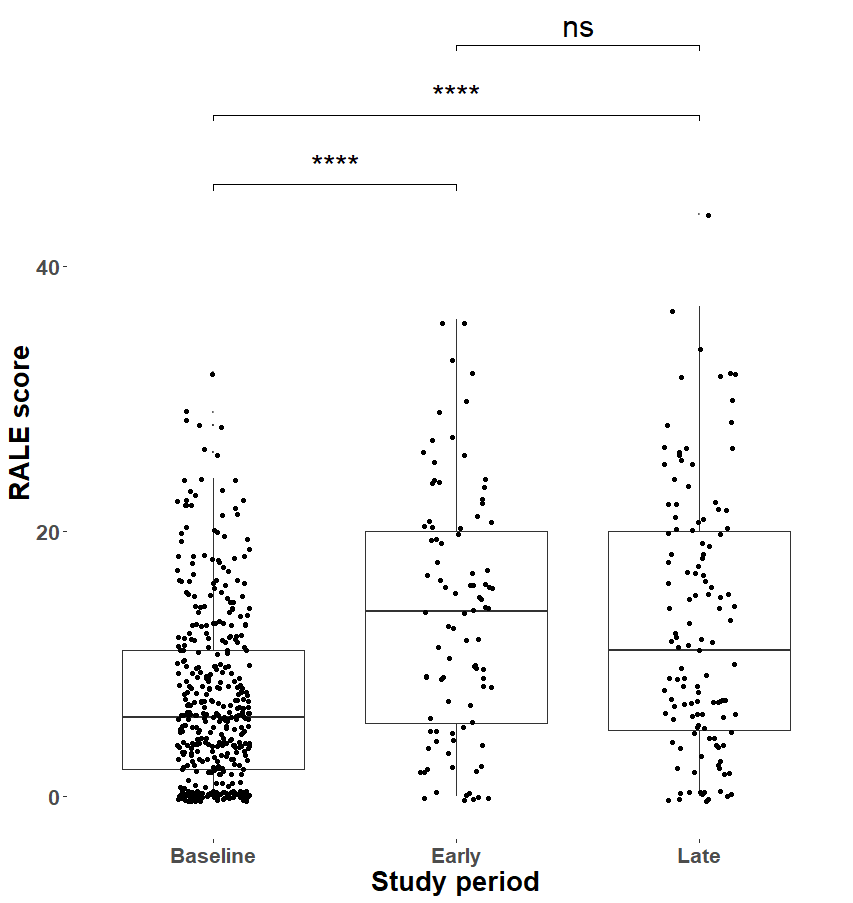

**Supplemental Table 1. Baseline characteristics of included subjects recording at time of ED evaluation stratified by RALE quartile.** P-values from nonparametric tests are shown in bold when significant (p<0.05). BMI: Body mass index; SBP: Systolic blood pressure; DBP: Diastolic blood pressure; S:F Ratio: Ratio of pulse oximetry oxygen saturation and fraction of inspired oxygen; WBC: white blood cell; Abs: absolute; NLR: Neutrophil-to-lymphocyte ratio; INR: international normalized ratio; BUN: Blood urea nitrogen; RALE Score: Radiographic assessment of lung edema score; OSA: Obstructive sleep apnea; DM: diabetes mellitus; CKD: chronic kidney disease; CLD: chronic liver disease; HTN: hypertension; CHF: congestive heart failure; HLD: hyperlipidemia.

| RALE Quartile | 1st | 2nd | 3rd | 4th | p |
| --- | --- | --- | --- | --- | --- |
| n | 376 | 78 | 216 | 200 |  |
| Male (%) | 160 (42.6) | 40 (51.3) | 114 (52.8) | 114 (57.0) | **0.005** |
| Age (median [IQR]) | 43.35 [31.80, 56.31] | 51.38 [39.14, 63.14] | 58.84 [48.76, 71.91] | 63.22 [53.31, 73.19] | **<0.001** |
| BMI (median [IQR]) | 29.98 [25.96, 34.72] | 32.29 [27.32, 37.79] | 30.55 [27.48, 35.88] | 30.70 [26.38, 36.69] | 0.101 |
| Symptom duration (median [IQR]) | 3.00 [2.00, 6.50] | 4.00 [2.00, 7.00] | 5.00 [2.00, 7.00] | 5.00 [2.00, 7.00] | **0.001** |
| SBP (median [IQR]) | 133.00 [122.25, 146.00] | 131.00 [121.25, 147.00] | 129.50 [116.00, 144.25] | 130.00 [116.00, 146.00] | 0.080 |
| DBP (median [IQR]) | 80.00 [71.25, 89.00] | 77.00 [67.50, 85.75] | 73.00 [65.00, 82.25] | 76.00 [65.00, 82.50] | **<0.001** |
| Pulse pressure (median [IQR]) | 53.00 [42.25, 64.00] | 54.50 [44.00, 69.00] | 54.50 [45.00, 69.00] | 56.00 [42.00, 68.00] | 0.218 |
| Respiratory rate (median [IQR]) | 18.00 [16.00, 20.00] | 18.00 [16.00, 20.00] | 18.00 [17.00, 20.00] | 20.00 [18.00, 24.00] | **<0.001** |
| Heart rate (median [IQR]) | 90.00 [79.50, 101.00] | 92.50 [79.25, 104.00] | 90.00 [76.00, 103.00] | 90.00 [80.00, 105.00] | 0.526 |
| Temperature (C) (median [IQR]) | 37.20 [36.80, 37.60] | 37.20 [36.90, 37.70] | 37.20 [36.80, 37.62] | 37.20 [36.70, 37.80] | 0.839 |
| SF Ratio (median [IQR]) | 466.67 [457.14, 476.19] | 457.14 [452.38, 466.67] | 457.14 [447.62, 466.67] | 447.62 [351.85, 461.90] | **<0.001** |
| Hemoglobin (median [IQR]) | 13.40 [12.40, 14.40] | 13.30 [12.20, 14.50] | 13.40 [11.90, 14.50] | 12.90 [11.30, 14.10] | **0.009** |
| Platelet count (median [IQR]) | 209.50 [168.25, 261.50] | 212.00 [157.00, 254.00] | 201.00 [158.00, 245.00] | 207.50 [166.25, 276.00] | 0.265 |
| WBC count (median [IQR]) | 5.34 [4.37, 6.76] | 5.08 [3.98, 6.22] | 5.68 [4.38, 7.35] | 6.86 [4.89, 9.12] | **<0.001** |
| Abs lymph (median [IQR]) | 1.29 [0.95, 1.78] | 1.19 [0.86, 1.57] | 1.04 [0.69, 1.47] | 0.88 [0.69, 1.29] | **<0.001** |
| Abs neutro (median [IQR]) | 3.29 [2.34, 4.56] | 3.13 [2.16, 4.47] | 4.02 [2.82, 5.31] | 5.17 [3.42, 7.17] | **<0.001** |
| NLR (median [IQR]) | 2.45 [1.61, 4.04] | 3.14 [1.89, 4.12] | 3.99 [2.65, 6.40] | 5.56 [3.36, 9.04] | **<0.001** |
| INR (median [IQR]) | 1.00 [1.00, 1.10] | 1.02 [1.00, 1.10] | 1.00 [1.00, 1.10] | 1.02 [1.00, 1.10] | 0.962 |
| Sodium (median [IQR]) | 138.00 [135.00, 139.00] | 136.50 [135.00, 138.00] | 137.00 [134.00, 139.00] | 136.00 [133.25, 138.00] | **<0.001** |
| BUN(median [IQR]) | 12.00 [8.25, 16.00] | 12.00 [9.00, 18.75] | 14.00 [10.00, 22.00] | 17.00 [12.00, 28.00] | **<0.001** |
| Creatinine (median [IQR]) | 0.89 [0.72, 1.10] | 0.98 [0.78, 1.18] | 0.96 [0.77, 1.23] | 1.02 [0.80, 1.47] | **<0.001** |
| BUN/Cr (median [IQR]) | 13.36 [10.21, 17.08] | 12.39 [9.19, 17.56] | 14.12 [11.49, 18.80] | 15.80 [11.93, 21.20] | **<0.001** |
| Potassium (median [IQR]) | 3.90 [3.70, 4.10] | 3.95 [3.60, 4.23] | 3.90 [3.60, 4.20] | 4.10 [3.70, 4.40] | **0.017** |
| Chloride (median [IQR]) | 101.00 [98.00, 104.00] | 99.00 [97.00, 102.00] | 99.00 [97.00, 102.00] | 98.00 [95.00, 101.00] | **<0.001** |
| Glucose (median [IQR]) | 102.00 [93.00, 119.75] | 109.00 [96.00, 128.50] | 117.00 [101.00, 150.00] | 121.00 [106.00, 156.50] | **<0.001** |
| Bicarbonate (median [IQR]) | 24.00 [23.00, 26.00] | 25.00 [22.00, 26.75] | 24.00 [22.00, 26.00] | 24.00 [21.00, 26.00] | 0.091 |
| Calcium (median [IQR]) | 9.10 [8.70, 9.30] | 8.90 [8.60, 9.20] | 8.90 [8.60, 9.20] | 8.85 [8.53, 9.10] | **<0.001** |
| RALE score (median [IQR]) | 0.00 [0.00, 0.00] | 2.00 [1.00, 2.00] | 5.00 [4.00, 6.00] | 12.00 [9.00, 16.00] | **<0.001** |
| History of OSA (%) | 21 ( 5.6) | 6 ( 7.7) | 25 (11.6) | 29 (14.5) | **0.003** |
| History of lung disease (%) | 72 (19.3) | 15 (19.2) | 48 (22.2) | 60 (30.0) | **0.027** |
| History of DM (%) | 53 (14.2) | 19 (24.4) | 56 (25.9) | 55 (27.5) | **<0.001** |
| History of immunosuppression (%) | 14 ( 3.7) | 4 ( 5.1) | 22 (10.2) | 13 ( 6.5) | **0.018** |
| History of CKD (%) | 15 ( 4.0) | 4 ( 5.1) | 24 (11.1) | 38 (19.0) | **<0.001** |
| History of CLD (%) | 3 ( 0.8) | 1 ( 1.3) | 6 ( 2.8) | 6 ( 3.0) | 0.179 |
| Hx of HTN(%) | 117 (31.3) | 36 (46.2) | 123 (56.9) | 124 (62.0) | **<0.001** |
| Hx of CHF (%) | 6 ( 1.6) | 4 ( 5.1) | 13 ( 6.0) | 32 (16.0) | **<0.001** |
| Hx of HLD(%) | 70 (18.6) | 24 (30.8) | 83 (38.4) | 103 (51.5) | **<0.001** |
| Hx of transplant (%) | 4 ( 1.1) | 0 ( 0.0) | 3 ( 1.4) | 6 ( 3.0) | 0.193 |
| Hx of tobacco use (%) | 124 (33.6) | 30 (38.5) | 81 (37.9) | 83 (41.7) | 0.277 |
| Hx of ethanol use (%) | 196 (53.3) | 31 (39.7) | 95 (44.8) | 74 (37.8) | **0.002** |
| 30-day mortality (%) | 2 ( 0.5) | 1 ( 1.3) | 29 (13.4) | 33 (16.5) | **<0.001** |
